# Sex differences in genetic liability to modifiable Alzheimer’s disease risk factors

**DOI:** 10.64898/2026.10.01.26364536

**Authors:** Rakshya U Sharma, Paulina Tolosa-Tort, Meri Okorie, Aadrita Chatterjee, Caroline Jonson, Kristine Yaffe, Michaël E Belloy, Shea J Andrews, ADGC and HABS-HD Study team

**Affiliations:** Department of Psychiatry and Behavioral Sciences, University of California, San Francisco, San Francisco, CA, USA; Department of Neurology and Weill Institute for Neurosciences, University of California, San Francisco, San Francisco, CA, USA; Department of Neurology, Washington University School of Medicine, St. Louis, MO, USA; NeuroGenomics and Informatics Center, Washington University School of Medicine, St. Louis, MO, USA; Edward and Pearl Fein Memory and Aging Center, University of California, San Francisco, San Francisco, CA, USA; DataTecnica LLC, Washington, DC USA; Center for Alzheimer’s and Related Dementias, National Institutes of Health, Bethesda, MD, USA

**Author notes:** Corresponding Author Shea J Andrews Assistant Professor Department of Psychiatry and Behavioral Sciences University of California, San Francisco Nancy Friend Pritzker Psychiatry Building 675 18th St.

**Keywords:** Alzheimer disease, polygenic risk score, modifiable risk factors, sex differences, genetic correlation, Mendelian randomization

## Abstract

**INTRODUCTION:** Alzheimer’s disease (AD) shows sex differences in risk, clinical progression, and biomarker profiles, but whether inherited genetic liability to modifiable risk factors contributes differently to AD susceptibility in females and males remains unclear. We evaluated whether genetic relationships between several modifiable dementia risk factors and AD differ between females and males.

**METHODS:** We integrated polygenic risk scores (PRSs) for modifiable risk factors, genome-wide genetic correlation, and two-sample Mendelian randomization (MR) analyses. PRS associations with AD diagnosis were evaluated in the Alzheimer’s Disease Genetics Consortium (n=27,751) and with AD plasma biomarkers in the Health and Aging Brain Study–Health Disparities (n=2,307-2,737, depending on the biomarker). Primary analyses evaluated female- and male-specific associations and compared sex-specific estimates.

**RESULTS:** Educational attainment and physical activity PRSs were associated with AD in both sexes. PRS sex differences were nominal for educational attainment, total cholesterol, and LDL cholesterol, but none remained significant after FDR correction. Genetic correlation showed FDR-significant sex differences for social isolation, physical activity, LDL cholesterol, and pulse pressure. MR identified FDR-significant associations within sex strata; higher HDL cholesterol was associated with lower AD risk in females, whereas higher total cholesterol and type 2 diabetes were associated with lower and higher AD risk, respectively, in males. Sensitivity analyses provided inconsistent support, and no sex difference in MR estimates remained significant after FDR correction. In HABS-HD, type 2 diabetes PRS was associated with higher NfL, BMI PRS with higher pTau_181_, and the composite PRS with higher NfL among males after FDR correction.

**DISCUSSION:** Evidence that modifiable dementia risk factors relate differently to AD by sex was limited and depended on the genetic measure examined. Sex differences in genome-wide genetic overlap were not consistently reflected in PRS or MR, indicating that shared genetic architecture does not necessarily translate into sex-specific inherited liability or causal effects

**RESEARCH IN CONTEXT:** *Systematic review:* We reviewed the published literature on modifiable dementia risk factors, Alzheimer’s disease (AD) genetics, and sex differences using PubMed and relevant reference lists. Previous studies support genetic relationships between several modifiable risk factors and AD, but systematic comparisons of female- and male-specific relationships across complementary genetic approaches remain limited.

*Interpretatio:* The strongest evidence of sex heterogeneity for modifiable risk factors and AD was observed in their shared genetic architectures, with FDR-significant female–male differences for social isolation, physical activity, LDL cholesterol, and pulse pressure. PRS and Mendelian randomization analyses provided less evidence of formal sex heterogeneity, suggesting that sex differences in shared genetic architecture do not necessarily imply sex-specific causal effects.

*Future directions:* Larger sex-stratified and ancestry-diverse GWAS integrated with longitudinal exposure and biomarker data are needed to determine whether these signals reflect sex-dependent biological mechanisms and whether they have implications for dementia prevention.

*Highlights:*

- Sex differences in AD-related genetic architecture were concentrated in behavioral, lipid, and vascular traits.
- Genetic correlations with AD differed between females and males for social isolation, physical activity, LDL cholesterol, and pulse pressure.
- PRS analyses implicated educational attainment and physical activity in AD in both sexes, but no PRS sex difference survived FDR correction.
- MR identified associations within sex strata but provided little robust evidence of sex-specific causal effects.

## 1. BACKGROUND

Sex differences in Alzheimer’s disease (AD) include a higher lifetime risk of developing AD and faster cognitive decline following diagnosis among women, as well as higher cerebrospinal fluid total tau levels in women and higher neurofilament light levels in men ^1^. Sex differences in AD risk may partly sex-specific relationships with modifiable cardiometabolic, behavioral, psychosocial, and sensory risk factors and differences in their underlying genetic architecture ^1,2^. A recent analysis identified polygenic gene-by-sex interactions across a range of complex traits, suggesting that their genetic architecture may differ between females and males ^3^. However, whether inherited liability to modifiable dementia risk factors contributes differently to AD susceptibility in females and males remains unclear. ^2,4^

Modifiable risk factors are central to dementia prevention, but their contribution to AD risk may differ between men and women ^5,6^. Approximately 45% of dementia cases can be attributed to 14 potentially modifiable risk factors across the life course, including lower education attainment, hearing loss, high LDL cholesterol, depression, traumatic brain injury, physical inactivity, diabetes, smoking, hypertension, obesity, excessive alcohol consumption, social isolation, air pollution, and vision loss ^7^. However, these risk factors may influence AD through pathways that are sex-specific, reflecting differences in exposure prevalence, timing, biological vulnerability, and downstream consequences for brain aging ^1,8,9^. Most prior work has focused on measured risk factors in observational studies, which may be affected by confounding, selection, and reverse causation. Evaluating inherited liability to these modifiable traits provides a complementary approach for determining whether genetic propensity toward dementia-relevant risk-factor pathways contributes to sex-specific AD susceptibility.

Genetic approaches can help clarify relationships between modifiable risk factors and AD because inherited genetic variation is generally less susceptible to confounding and reverse causation ^4,10^. Polygenic risk scores (PRS) can estimate inherited liability to modifiable traits and test whether this liability is associated with AD risk or AD-related endophenotypes. Genetic correlation can assess whether modifiable traits and AD share common variant genetic architecture, while Mendelian randomization (MR) can assess whether genetic evidence is consistent with potential causal effects under instrumental-variable assumptions ^11^. Together with conventional epidemiological studies, triangulation across complementary genetic approaches with different assumptions and sources of bias can provide more complete assessment of sex-specific genetic relationships than any single method alone ^12^. However, it remains unclear whether evidence across complementary genetic approaches converges on sex-specific relationships between inherited liability to modifiable dementia risk and AD susceptibility ^2,4,5^.

In this study, we evaluated whether genetic relationships between modifiable dementia risk factors and AD differ between females and males. We integrated individual level PRS analyses with sex-stratified genome-wide genetic correlation and MR to evaluate associations with AD susceptibility, shared genetic architecture and evidence consistent with potential causal relationships. We compared female and male specific estimates across these approaches. Phenotypic validation of trait-specific PRS and associations with plasma AD biomarkers in Health and Aging Brain Study-Health Disparities cohort were evaluated as complementary analyses.

## 2. METHODS

### 2.1. Study Overview

We evaluated whether genetic liability to modifiable dementia risk factors—including diabetes, total cholesterol, HDL cholesterol, LDL cholesterol, triglycerides, BMI, pulse pressure, diastolic blood pressure, systolic blood pressure, education, depression, alcohol consumption, smoking, physical activity, hearing loss, vision loss and social isolation—was associated with AD and whether these relationships differed between females and males. Analyses integrated PRS, genetic correlation (r_g_), and MR (see Supplementary Figure S1). The primary analyses evaluated female- and male-specific associations with AD and tested differences between sex-specific estimates. Combined-sex analyses were performed as supporting analyses. PRS validation and association with plasma biomarkers of AD pathology were evaluated in the Health and Aging Brain Study-Health Disparities (HABS-HD) cohort.

### 2.2 Study populations

#### 2.2.1 Alzheimer’s Disease Genetics Consortium (ADGC)

The Alzheimer’s Disease Genetics Consortium (ADGC) is a large-scale collaborative initiative comprising case-control, family-based, and population-based cohorts across the United States, established to identify genetic variants associated with the risk of late-onset AD ^13^. AD cases were defined using neuropathological confirmation or cohort-specific clinical diagnostic criteria, whereas controls were cognitively unimpaired individuals without mild cognitive impairment (MCI) or AD based on cohort-specific cognitive evaluation ^14^. Written informed consent was obtained from study participants or, for those with substantial cognitive impairment, from a caregiver, legal guardian, or other proxy, and the study protocols for all populations were reviewed and approved by the appropriate institutional review boards (IRBs).

Genotype data underwent pre-imputation, imputation, and post-imputation quality control at both the sample and variant levels (see Supplementary Methods) ^14–16^. Participants were required to have AD case-control status, age, sex, *APOE ε4* status, the first four genetic ancestry principal components, and non-missing polygenic risk scores (PRSs) for all evaluated modifiable risk factors. The final analytic sample comprised 27,751 participants (AD = 10,532, Controls = 17,219, male = 10,959; female = 16,792).

#### 2.2.2 Health and Aging Brain Study – Health Disparities (HABS-HD)

The Health and Aging Brain Study-Health Disparities (HABS-HD) is a community-based cohort designed to investigate cognitive aging, AD and related dementias, associated biomarkers, and health disparities in a racially and ethnically diverse aging population ^17^. HABS-HD recruited Black, Hispanic, and non-Hispanic White older adults through the University of North Texas Health Science Center in Fort Worth, Texas; eligibility and exclusion criteria have been described previously ^17,18^.

For the present study, we used baseline clinical data available in HABS-HD data release 7. Self-reported questionnaires and clinical assessments were used to characterize modifiable dementia risk factors (see Supplementary Methods; Table 1). Cognitive impairment was defined as the presence of mild cognitive impairment or dementia. Plasma biomarkers of AD pathology, including pTau_181_, pTau_217_, Aβ_42_/Aβ_40_, NfL, and total tau have been described previously ^19^. The final HABS-HD analytic sample comprised 3,207 participants. Detailed phenotype definitions, biomarker processing and inclusion criteria are provided in Supplementary Methods.

**Table 1.** Characteristics of HABS-HD and ADGC analytic samples, stratified by sex.

| Characteristic | HABS-HD |  |  | ADGC |  |  |
| --- | --- | --- | --- | --- | --- | --- |
|  | Female<br>N = 1981<sup>1</sup> | Male<br>N = 1226<sup>1</sup> | P value<sup>2</sup> | Female<br>N = 16792 | Male<br>N = 10959 | P value |
| Age | 65.08 (8.63) | 66.36 (8.54) | <0.001 | 74.85 (8.87) | 74.73 (8.49) | 0.2 |
| Race/ethnicity |  |  | <0.001 |  |  |  |
| Black | 24.7% | 23.5% |  |  |  |  |
| Hispanic | 34.8% | 42.1% |  |  |  |  |
| NHW | 40.5% | 34.4% |  |  |  |  |
| Education (years) | 13.08 (4.24) | 13.72 (4.43) | <0.001 |  |  |  |
| BMI | 30.75 (7.13) | 30.19 (5.87) | 0.016 |  |  |  |
| Total cholesterol (mg/dL) | 190.09 (40.09) | 168.50 (40.18) | <0.001 |  |  |  |
| LDL cholesterol (mg/dL) | 107.40 (33.61) | 96.95 (33.86) | <0.001 |  |  |  |
| HDL cholesterol (mg/dL) | 59.47 (16.01) | 48.47 (12.91) | <0.001 |  |  |  |
| Triglycerides (mg/dL) | 124.56 (72.50) | 128.25 (78.15) | 0.2 |  |  |  |
| HbA1c | 5.97 (1.30) | 6.04 (1.34) | 0.2 |  |  |  |
| Diastolic BP (mmHg) | 82.30 (10.52) | 84.68 (10.78) | <0.001 |  |  |  |
| Systolic BP (mmHg) | 133.44 (19.11) | 138.86 (17.73) | <0.001 |  |  |  |
| Pulse pressure (mmHg) | 51.14 (14.85) | 54.18 (13.24) | <0.001 |  |  |  |
| GDS score | 5.92 (5.82) | 5.30 (5.35) | 0.002 |  |  |  |
| Hypertension | 62.4% | 71.0% | <0.001 |  |  |  |
| Diabetes | 23.4% | 26.8% | 0.033 |  |  |  |
| Clinical depression | 38.0% | 25.5% | <0.001 |  |  |  |
| Alcohol use score | 1.38 (1.74) | 2.64 (2.95) | <0.001 |  |  |  |
| Social support score | 40.99 (6.31) | 40.57 (6.34) | 0.066 |  |  |  |
| History of TBI | 1.4% | 3.1% | 0.001 |  |  |  |
| Smoker | 30.0% | 47.5% | <0.001 |  |  |  |
| Physically active | 10.5% | 18.2% | <0.001 |  |  |  |
| Cognitive status |  |  | <0.001 |  |  |  |
| Cognitively unimpaired | 77.3% | 66.3% |  |  |  |  |
| MCI/dementia | 22.7% | 33.7% |  |  |  |  |
| APOE ε4 carrier | 28.7% | 28.5% | >0.9 | 38.3% | 38.4% | >0.9 |
| pTau<sub>181</sub> | -0.11 (0.92) | 0.17 (1.10) | <0.001 |  |  |  |
| pTau<sub>217</sub> | -0.08 (0.98) | 0.13 (1.02) | <0.001 |  |  |  |
| Aβ<sub>42</sub>/Aβ<sub>40</sub> | 0.00 (1.00) | 0.02 (1.00) | 0.6 |  |  |  |
| NfL | -0.02 (1.00) | 0.03 (1.00) | 0.2 |  |  |  |
| Total tau | 0.17 (1.00) | -0.28 (0.94) | <0.001 |  |  |  |
| ADGC case-control status§ |  |  |  |  |  | <0.001 |
| Cognitively unimpaired |  |  |  | 63.2% | 60.3% |  |
| Alzheimer's disease |  |  |  | 36.8% | 39.7% |  |

<sup>1</sup>Mean (SD); %<sup>2</sup>Welch Two Sample t-test; Pearson's Chi-squared test
Note: Values are mean (SD) for continuous variables and percentages for categorical variables. †In HABS-HD, cognitive impairment includes mild cognitive impairment (MCI) and dementia. #Physically active reflects the percentage classified in the high-score category. §In ADGC, case-control status was defined as cognitively unimpaired or Alzheimer's disease according to the harmonized ADGC phenotype. APOE ε4 carrier and smoker rows show the percentage in the positive category.

### 2.3 GWAS Summary Statistics

We used GWAS summary statistics for PRS construction, genetic correlation, and MR analyses (see Supplementary Methods and Table S1) for specific GWAS sources, phenotype definitions, sample sizes, and corresponding analytical applications. For PRS construction, we used the largest available published GWAS of each modifiable risk factor (samples ranging from 377,234 to 3,383,199 participants) ^20–30^. For genetic correlation and MR analyses, we used combined-sex and sex-stratified AD GWAS summary statistics combining data from ADGC, ADSP, UK Biobank, and FinnGen ^31^, together with female- and male-specific GWAS summary statistics for modifiable risk factors from the Neale Lab UK Biobank GWAS ^32^.

### 2.4 Polygenic Risk Scores

Trait-specific PRS were constructed using PRS-CS-Auto which applies a Bayesian continuous-shrinkage prior while accounting for linkage disequilibrium ^33^. The *APOE* region was excluded to minimize associations driven by the major AD susceptibility locus rather than genome-wide liability to the modifiable risk factor ^3^. PRS were ancestry standardized using *pgsc_calc* ^34^ and standardized to represent one-standard deviation increase in genetic liability.

We additionally constructed a multi-trait composite PRS using a stratified 70% training and 30% held-out test framework in ADGC ^35^. Trait-specific PRS coefficients estimated in the training data were used to construct the composite score, which was evaluated for association with AD in the held-out test set. Composite models were constructed separately for the combined, female and male samples. Ridge, elastic-net, and leave-one-PRS-out models were evaluated as sensitivity analyses (see Supplementary Methods).

### 2.5 Statistical Analysis

#### 2.5.1 Cohort Description

Participant characteristics were summarized separately by sex for ADGC and for HABS-HD. Differences between females and males were assessed using two-sample *t-*tests for continuous variable and Pearson’s chi-square test for categorical variables.

#### 2.5.2 PRS Association Analyses

In HABS-HD, trait-specific PRS were first evaluated against their corresponding measured phenotypes to assess phenotypic concordance. Associations of trait specific composite PRS with plasma AD biomarkers were subsequently evaluated using linear regression. Models were adjusted for age, *APOE ε4* status, race/ethnicity, first four principal components, and sex where applicable (see Supplementary Methods).

In ADGC, we evaluated the association between each trait-specific PRS and AD case-control status using logistic regression adjusting for age, *APOE ε4* status, sex, and the first four genetic principal components. To assess the sex-specific associations, we repeated the logistic regression analyses separately among female and male participants.

#### 2.5.3 Genetic Correlations

Genome-wide genetic correlations between modifiable dementia risk factors and AD were estimated using GNOVA (see Supplementary methods) ^36^. Analyses were conducted separately using combined-sex, female-specific, and male-specific GWAS summary statistics. The primary analyses used AD GWAS summary statistics combining ADGC, ADSP, UK Biobank, and FinnGen. Sensitivity analyses repeated the genetic-correlation analyses using alternative AD GWAS meta-analyses excluding either UK Biobank or FinnGen.

#### 2.5.4 Mendelian Randomization

We performed two-sample Mendelian randomization (MR) to assess potential causal associations between genetically predicted modifiable risk factors and AD (see Supplementary Methods). Independent genome-wide significant variants (*P* < 5 × 10^-8^; *r*² < 0.001) were selected as genetic instruments, and the *APOE* region was excluded. The primary MR analysis used a fixed effects inverse-variance weighted method and required at least three retained genetic instruments. Sensitivity analyses included weighted median, weighted mode, and MR-Egger regression. Heterogeneity among genetic instruments was assessed using Cochran’s Q statistic, and directional horizontal pleiotropy was evaluated using the MR-Egger intercept test. MR-PRESSO was additionally used to assess global heterogeneity and identify potential outlier instruments; where outliers were detected, MR analyses and sensitivity tests were repeated after their removal.

#### 2.5.5 Statistical significance and sex-difference testing

Differences between female-and male-specific effect estimates were evaluated using a Z-test, calculated as

*Z* = (*β_F_* - *β_M_*) / √((*SE_F_*)^2^ + *SE_M_*)^2^),

where *β_F_* and *β_M_* are the female and male effect estimates respectively and *SE_F_* and *SE_M_* are their corresponding standard errors from the female and male stratified models, respectively ^37^. Two-sided P-values were calculated from Z statistics. Z-tests were applied to sex-specific estimates from PRS, genetic correlation and MR analyses.

Within each analysis (PRS, r_g_, MR), P-values were adjusted for multiple testing using the Benjamin-Hochberg false discovery rate (FDR) method. Association for female-male differences with *P* < 0.05 but FDR-adjusted *P ≥* 0.05 were considered nominally significant, whereas those with FDR-adjusted *P <* 0.05 were considered statistically significant.

## 3. RESULTS

### 3.1. Cohort description of ADGC and HABS-HD

The HABS-HD analytic sample included 3,207 participants (1,226 males and 1,981 females), and the ADGC analytic sample included 27,751 participants (16,792 females and 10,959 males). In ADGC, 38% of participants were AD cases, and 80% were of European ancestry (Table 1).

### 3.2 PRS validation and associations with AD and plasma biomarkers

Trait-specific PRSs were evaluated against their corresponding phenotypes in HABS-HD to assess whether the scores captured their intended phenotypic domains (Supplementary Figure S2; Supplementary Table S2). All evaluated PRSs were significantly associated with their corresponding phenotypes after FDR correction and effect directions were consistent with the corresponding trait definitions.

In the combined sex analyses in ADGC, higher educational attainment and diastolic blood pressure PRSs were associated with lower odds of AD, whereas higher physical activity PRS was associated with greater odds of AD (Figure 1; Supplementary Table S3). A higher composite PRS was associated with higher odds of AD, with similar effect estimates across logistic, ridge, and elastic-net weighting approaches (Supplementary Table S4). In Leave-one-PRS-out analyses, removal of educational attainment PRS produced the greatest attenuation of the composite association, indicating that educational attainment contributed substantially to the composite PRS signal (Supplementary Figure S3).

**Figure 1:**
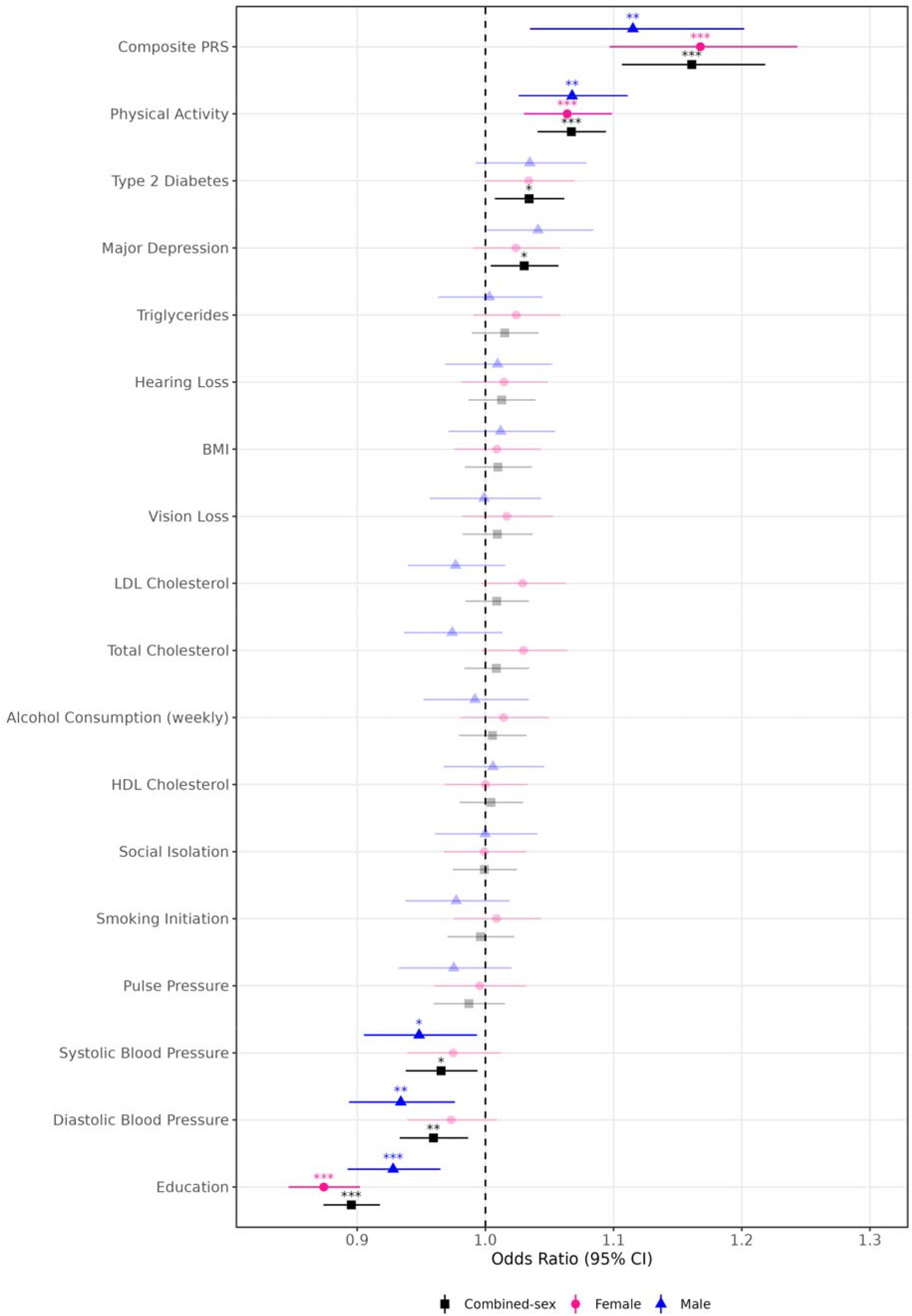
Associations of individual and composite polygenic risk scores with Alzheimer’s disease in ADGC, combined-sex and stratified by sex. Forest plot showing odds ratios (ORs) and 95% confidence intervals for associations between each modifiable risk factor polygenic risk score and Alzheimer’s disease in the Alzheimer’s Disease Genetics Consortium (ADGC). Black squares indicate the combined-sex ADGC estimate, pink circles indicate female-specific estimates, and blue triangles indicate male-specific estimates. The composite PRS is shown in the top row, and individual PRSs are shown below. The dashed vertical line marks the null value (OR = 1). Significance symbols denote false discovery rate (FDR)-adjusted associations within each stratum: *FDR < 0.05, **FDR < 0.01, ***FDR < 0.001.

We evaluated associations between trait-specific PRSs and plasma AD biomarkers in HABS-HD combined-sex analyses. Higher type 2 diabetes PRS was associated with higher Nfl, and higher BMI PRS was associated with higher pTau_181_, no other associations remained significant after FDR correction (Figure 2; Supplementary Table S5).

**Figure 2:**
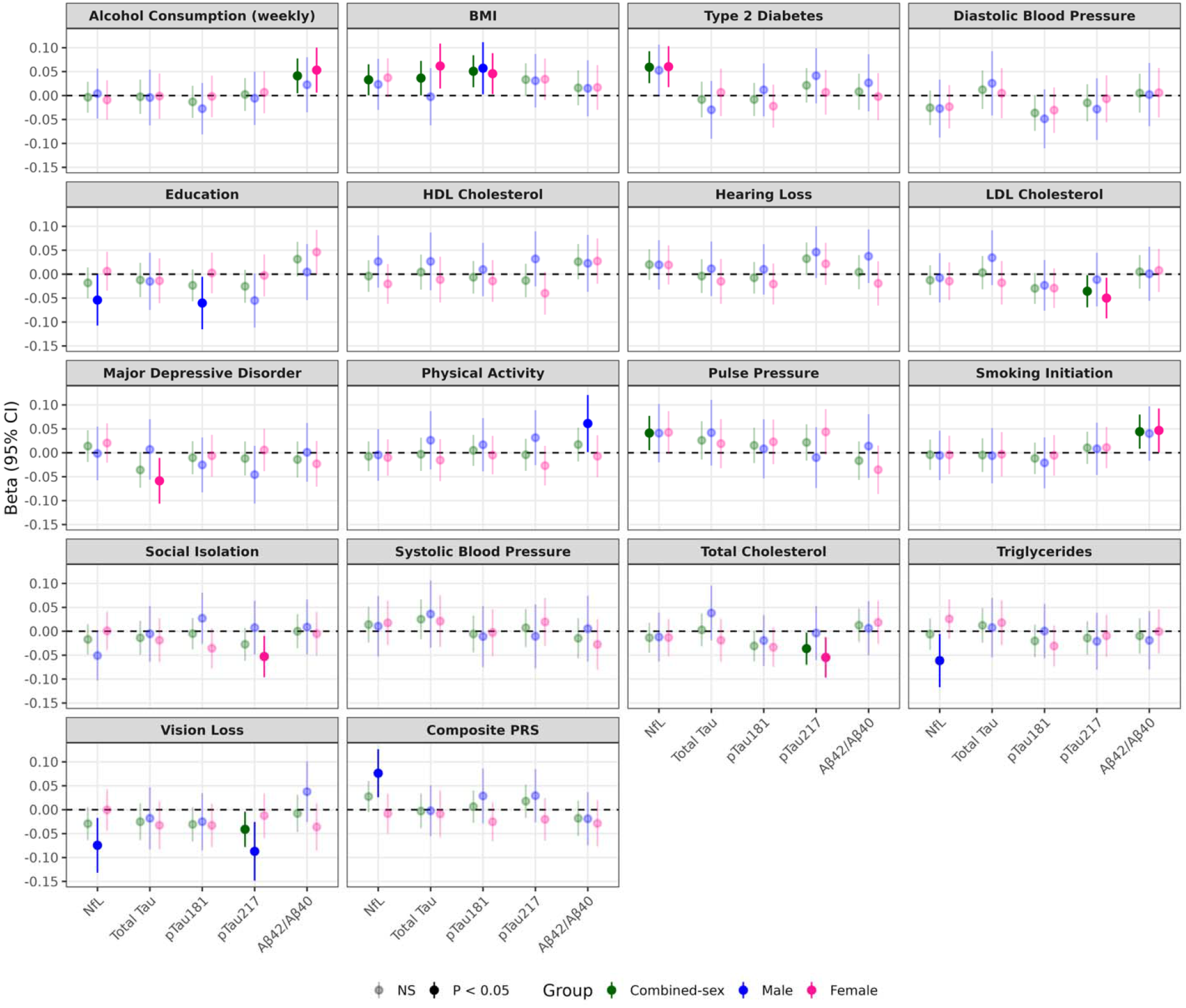
Associations of composite and individual polygenic risk scores with plasma endophenotypes in HABS-HD. Point estimates and 95% confidence intervals are shown for associations with NfL, total tau, *pTau_181_, pTau_217_, and A*β*_42_/A*β*_40_*. Facets display results for each individual PRS, with the composite PRS shown in the final panel. Models were adjusted for age, *APOE* ε*4* status, race/ethnicity, and genetic principal components 1–4; sex was additionally included in the combined-sex model and omitted in sex-stratified analyses. Colors indicate the analysis group: combined-sex, males, and females. Filled points indicate P < 0.05 before multiple-testing correction. Associations surviving false discovery rate correction were observed for type 2 diabetes PRS with NfL in the combined-sex, composite PRS with NfL in males, and BMI PRS with *pTau_181_* in the combined-sex. Full nominal and false discovery rate-adjusted p-values are reported in Supplementary Table S5.

### 3.3 Sex-specific genetic associations with AD

Sex-stratified analyses across PRS, genome-wide genetic correlation, and MR identified several genetic relationships between modifiable risk factors and AD (Figures 1, 3 and 4; Table 2; Supplementary Tables S6–S9). Female–male comparisons were used to distinguish statistical evidence of sex heterogeneity from associations observed within individual sex strata. Complete individual PRS, genetic-correlation and MR results are reported in Supplementary Tables S6, S7, and S8, respectively, and female-male comparison tests are reported in Supplementary Table S9. In sex-stratified analyses, the composite PRS was associated with higher NfL among males. No other PRS-biomarker association remained significant after FDR correction.

**Figure 3:**
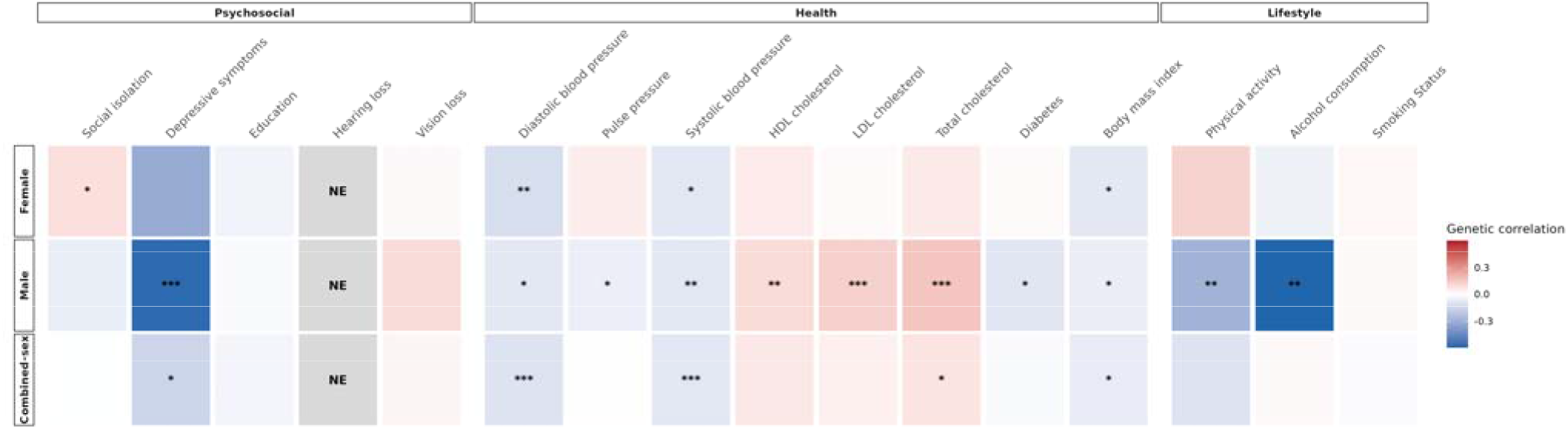
Genetic correlations between modifiable dementia risk factors and Alzheimer’s disease using ADGC, ADSP, UK Biobank and FinnGen. Heatmap showing GNOVA genome-wide genetic correlations between selected Lancet Commission modifiable dementia risk factors and Alzheimer’s disease, using ADGC, ADSP, UK Biobank and FinnGen as the primary Alzheimer’s disease outcome dataset. Columns represent modifiable-risk-factor traits grouped into psychosocial, health, and lifestyle domains. Rows show female-specific, male-specific, and combined Alzheimer’s disease GWAS analyses. Color indicates the direction and magnitude of the genetic correlation, with red representing positive genetic correlation and blue representing inverse genetic correlation. Asterisks indicate associations surviving FDR correction within each analysis stratum (*q < 0.05, **q < 0.01, ***q < 0.001) and NE indicated Non Estimable. Complete nominal and FDR-adjusted P values are provided in Supplementary Table S7.

**Figure 4.**
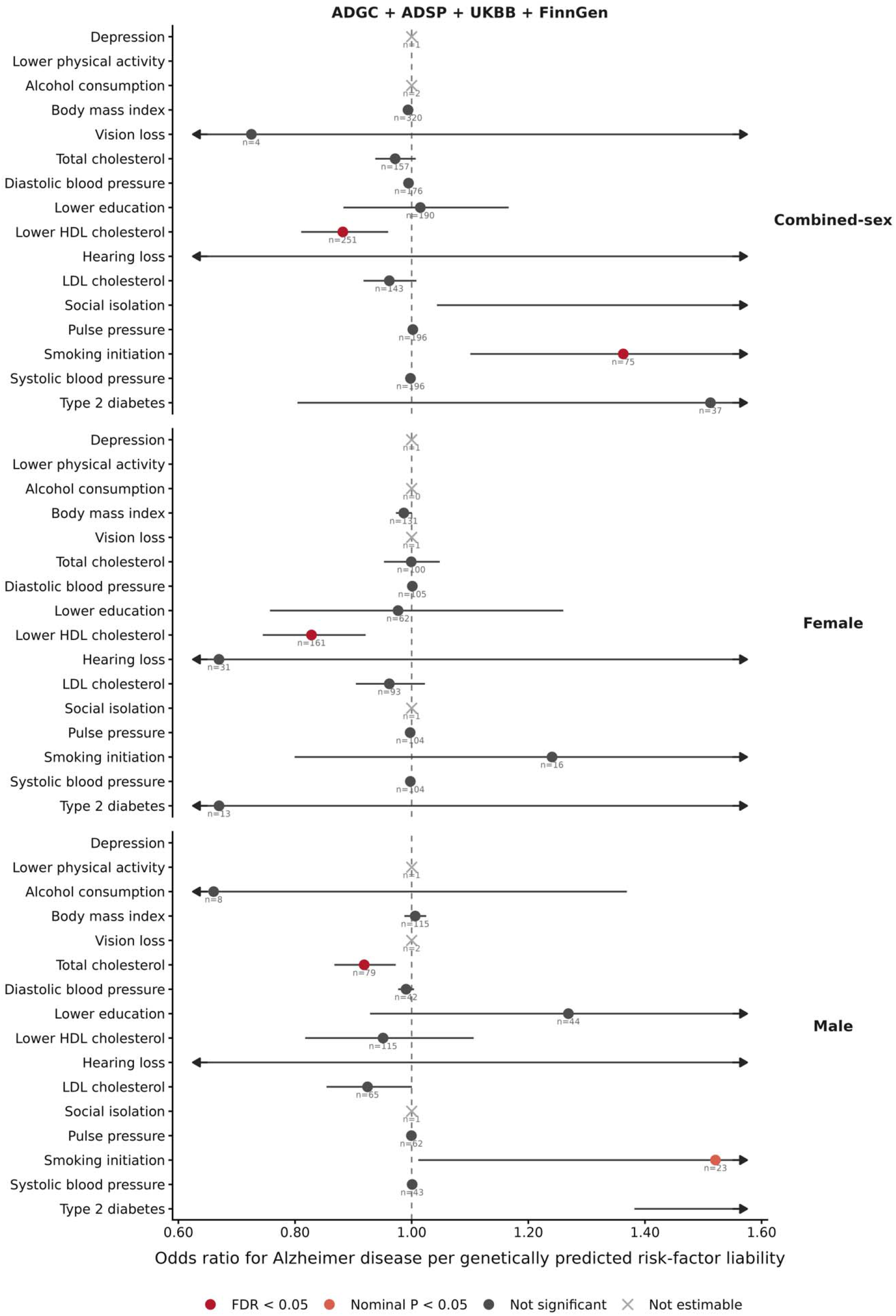
Mendelian randomization estimates for modifiable dementia risk factors and Alzheimer’s disease. Forest plot showing inverse-variance-weighted (IVW) two-sample Mendelian randomization estimates for associations between genetically predicted modifiable dementia risk factors and Alzheimer’s disease (AD) using the combined ADGC, ADSP, UK Biobank, and FinnGen AD GWAS outcome. Results are presented for combined-sex, female-specific, and male-specific analyses. Points indicate IVW effect estimates on the log-odds scale and horizontal lines indicate 95% confidence intervals; the dashed vertical line denotes the null value of 0. Positive estimates indicate higher genetically predicted risk-factor liability associated with higher odds of AD, whereas negative estimates indicate lower odds of AD. Orange points denote nominally significant associations (P < 0.05) and blue points denote P ≥ 0.05. Numbers adjacent to estimates indicate the number of genetic instruments included in each MR analysis. Crosses indicate exposure–outcome pairs excluded because fewer than three MR-ready genetic instruments were available. Exposure directions were harmonized to reflect risk-oriented liability where applicable, including lower educational attainment, lower HDL cholesterol, and lower physical activity.

**Table 2.** Sex-specific genetic associations of modifiable dementia risk factors with Alzheimer’s disease.

| Risk factor | Polygenic risk score |  |  | Genetic correlation |  |  | Mendelian randomization |  |  |
| --- | --- | --- | --- | --- | --- | --- | --- | --- | --- |
|  | Female | Male | q | Female | Male | q | Female | Male | q |
| Education | 1.14 (1.11, 1.18);<br>q=2.04e-15 | 1.08 (1.04, 1.12);<br>q=0.003 | 0.242 | 0.04; q=0.199 | 0.02; q=0.517 | 0.573 | -0.02 (-0.28, 0.23);<br>q=0.941 | 0.24 (-0.07, 0.55);<br>q=0.324 | 0.372 |
| Alcohol consumption | 1.01 (0.98, 1.05);<br>q=0.590 | 0.99 (0.95, 1.03);<br>q=0.913 | 0.829 | -0.05; q=0.788 | -0.72; q=0.009 | 0.053 | NA | -0.41 (-1.14, 0.31);<br>q=0.454 | NA |
| BMI | 1.01 (0.98, 1.04);<br>q=0.748 | 1.01 (0.97, 1.05);<br>q=0.884 | 0.977 | -0.08; q=0.039 | -0.06; q=0.023 | 0.648 | -0.01 (-0.03, 0.00);<br>q=0.287 | 0.01 (-0.01, 0.02);<br>q=0.631 | 0.353 |
| Type 2 diabetes | 1.03 (1.00, 1.07);<br>q=0.270 | 1.03 (0.99, 1.08);<br>q=0.314 | 0.977 | 0.01; q=0.788 | -0.09; q=0.015 | 0.106 | -0.40 (-1.88, 1.08);<br>q=0.937 | 1.12 (0.32, 1.91);<br>q=0.035 | 0.353 |
| Systolic blood pressure | 0.97 (0.94, 1.01);<br>q=0.342 | 0.95 (0.91, 0.99);<br>q=0.107 | 0.829 | -0.09; q=0.029 | -0.09; q=0.009 | 0.992 | -0.00 (-0.01, 0.00);<br>q=0.735 | 0.00 (-0.01, 0.01);<br>q=0.900 | 0.623 |
| Diastolic blood pressure | 0.97 (0.94, 1.01);<br>q=0.342 | 0.93 (0.89, 0.98);<br>q=0.014 | 0.663 | -0.12; q=0.007 | -0.08; q=0.015 | 0.573 | 0.00 (-0.01, 0.01);<br>q=0.941 | -0.01 (-0.02, 0.00);<br>q=0.333 | 0.372 |
| Pulse pressure | 0.99 (0.96, 1.03);<br>q=0.912 | 0.97 (0.93, 1.02);<br>q=0.474 | 0.829 | 0.05; q=0.199 | -0.06; q=0.048 | 0.039 | -0.00 (-0.01, 0.00);<br>q=0.735 | -0.00 (-0.01, 0.01);<br>q=0.900 | 0.660 |
| HDL cholesterol | 1.00 (0.97, 1.03);<br>q=0.999 | 0.99 (0.96, 1.03);<br>q=0.931 | 0.977 | -0.05; q=0.199 | -0.10; q=0.009 | 0.573 | -0.19 (-0.29, -0.08);<br>q=0.006 | -0.05 (-0.20, 0.10);<br>q=0.631 | 0.372 |
| LDL cholesterol | 1.03 (1.00, 1.06);<br>q=0.270 | 0.98 (0.94, 1.02);<br>q=0.474 | 0.242 | 0.01; q=0.776 | 0.13; q=7.04e-04 | 0.039 | -0.04 (-0.10, 0.02);<br>q=0.735 | -0.08 (-0.16, 0.00);<br>q=0.153 | 0.623 |
| Total cholesterol | 1.03 (1.00, 1.06);<br>q=0.270 | 0.97 (0.94, 1.01);<br>q=0.468 | 0.242 | 0.06; q=0.139 | 0.16; q=5.75e-05 | 0.075 | -0.00 (-0.05, 0.05);<br>q=0.974 | -0.08 (-0.14, -0.03);<br>q=0.035 | 0.297 |
| Triglycerides | 1.02 (0.99, 1.06);<br>q=0.342 | 1.00 (0.96, 1.04);<br>q=0.992 | 0.829 | NA | NA | NA | NA | NA | NA |
| Depression | 1.02 (0.99, 1.06);<br>q=0.342 | 1.04 (1.00, 1.08);<br>q=0.182 | 0.829 | -0.32; q=0.060 | -0.58; q=1.76e-04 | 0.317 | NA | NA | NA |
| Social isolation | 1.00 (0.97, 1.03);<br>q=0.999 | 1.00 (0.96, 1.04);<br>q=0.992 | 0.977 | 0.09; q=0.039 | -0.06; q=0.170 | 0.039 | NA | NA | NA |
| Physical activity | 0.94 (0.91, 0.97);<br>q=0.001 | 0.94 (0.90, 0.97);<br>q=0.011 | 0.977 | -0.12; q=0.474 | 0.28; q=0.008 | 0.039 | NA | NA | NA |
| Smoking | 1.01 (0.97, 1.04);<br>q=0.748 | 0.98 (0.94, 1.02);<br>q=0.474 | 0.829 | 0.02; q=0.474 | 0.02; q=0.559 | 0.920 | 0.22 (-0.22, 0.66);<br>q=0.735 | 0.42 (0.01, 0.83);<br>q=0.153 | 0.623 |
| Hearing loss | 1.01 (0.98, 1.05);<br>q=0.590 | 1.01 (0.97, 1.05);<br>q=0.913 | 0.977 | Unstable | Unstable | NA | -0.40 (-3.99, 3.18);<br>q=0.941 | 0.57 (-1.01, 2.15);<br>q=0.631 | 0.660 |
| Vision loss | 1.02 (0.98, 1.05);<br>q=0.590 | 1.00 (0.96, 1.04);<br>q=0.992 | 0.829 | 0.02; q=0.810 | 0.10; q=0.293 | 0.573 | NA | NA | NA |

Note. In each analysis block, the standalone q column denotes the FDR-adjusted P value for the formal female–male sex-difference Z-test. Within the female and male estimate cells, q denotes the FDR-adjusted P value for that sex-specific association. PRS values are ORs (95% CI), genetic-correlation values are $r_g$ estimates (95% CI), and MR values are IVW beta estimates (95% CI). PRS estimates represent associations per one-standard-deviation increase in the phenotype defined by the source GWAS. MR exposures were oriented toward greater dementia-risk liability where indicated. A significant estimate in one sex and not the other does not by itself establish a sex difference.

#### 3.3.1 Educational attainment and physical activity

Higher educational attainment PRS was associated with lower odds of AD in both females and males, with a larger inverse effect estimate among females (Figure 1; Table 2; Supplementary Table S6). The female–male difference was nominally significant but did not survive FDR correction (see Supplementary Figure S4).

Genome-wide genetic correlation analyses also supported shared genetic architecture between educational attainment and AD, although evidence for sex heterogeneity depended on the composition of the AD GWAS (Table 2; Figure 3; Supplementary Table S7). In sensitivity analyses excluding UK Biobank, evidence for a female–male difference was stronger than in the primary AD meta-analysis (Supplementary Figure S5; Supplementary Table S7). MR estimates varied across AD GWAS definitions and sensitivity analyses, providing limited evidence for a consistent sex-specific causal effect (Table 2; Supplementary Figure S6; Supplementary Table S8).

Higher physical activity PRS was associated with higher odds of AD in both females and males, but the female-male difference did not survive FDR correction (Figure 1; Table 2; Supplementary Table S6). In contrast, genetic correlation demonstrated an FDR-significant female–male difference, with a positive genetic correlation in females (*r_g_* = 0.12) and a negative genetic correlation in males (*r_g_* = -0.28; P_FDR_ = 0.039) (Figure 3; Table 2; Supplementary Tables S7 and S9). MR analyses did not provide corresponding evidence for a robust sex-specific causal effect (Table 2; Supplementary Table S8).

#### 3.3.2 Lipid traits

Sex-dependent patterns were observed for several lipid traits. PRS effect estimates for both LDL and total cholesterol differed nominally between females and males, although neither female–male comparison remained significant after FDR correction (Figure 1; Table 2; Supplementary Table S6).

In genetic correlation analyses, LDL cholesterol showed an FDR-significant female–male difference. The genetic correlation with AD was positive among males (*r_g_* = 0.13) but near zero among females (*r_g_* = 0.01; P_FDR_ for sex difference = 0.039) (Figure 3; Table 2; Supplementary Tables S7 and S9). In the primary analyses, HDL and total cholesterol also showed associations within individual sex strata, but their female-male difference did not remain significant after FDR correction.

MR identified sex-specific associations for HDL and total cholesterol in the primary analyses, but these findings were not accompanied by FDR-significant female-male differences. Evidence from sensitivity analyses varied across lipid traits and AD GWAS definitions, providing limited support for consistent sex heterogeneity in the causal effect (Table 2; Supplementary Tables S8 and S9).

#### 3.3.3 Blood Pressure and Cardiometabolic traits

Blood-pressure traits showed several sex-specific associations. Higher diastolic blood pressure PRS was associated with lower odds of AD among males, whereas the association was not significant among females; however, the female–male difference was non-significant (Figure 1; Table 2; Supplementary Tables S6 and S9).

In genetic correlation analyses, pulse pressure showed a negative genetic correlation with AD among males, with an FDR-significant female-male difference (Figure 3; Table 2; Supplementary Tables S7 and S9). Systolic and diastolic blood pressure showed additional genetic associations with AD within sex strata but not significant female–male difference in the primary analysis. MR analyses did not provide evidence of sex heterogeneity for blood-pressure traits in the primary analyses (Table 2; Supplementary Tables S8 and S9).

For BMI and type 2 diabetes, were inconsistent across PRS, genetic correlation, and MR analyses. Neither trait showed an FDR-significant female-male difference in the primary analyses, providing limited evidence for sex heterogeneity in the genetic relationships with AD (Table 2; Supplementary Tables S6–S9).

#### 3.3.4 Social Isolation and other modifiable risk factors

Social isolation showed evidence of sex-specific shared genetic architecture with AD. In the primary AD meta-analysis, social isolation showed a positive genetic correlation with AD among females and a negative genetic correlation among males, with an FDR-significant female-male difference (Figure 3; Table 2; Supplementary Tables S7 and S9).

For the remaining behavioral, psychiatric, and sensory risk factors, evidence for sex heterogeneity was limited (Table 2; Supplementary Tables S6–S9). Although associations were observed within individual female or male strata for some traits, formal female–male comparisons did not remain significant after FDR correction.

## 4. DISCUSSION

We found that the genetic relationships between modifiable dementia risk factors and AD are not uniformly shared between females and males. The clearest evidence of sex heterogeneity emerged from genetic correlations, with social isolation, physical activity, LDL cholesterol, and pulse pressure showing FDR-significant female-male differences. In contrast, no PRS female-male comparison survived FDR correction, and MR provided limited evidence for consistent sex heterogeneity. These findings suggest that sex differences may be more apparent in the shared genetic architecture linking selected behavioral, lipid, and vascular pathways to AD than in individual-level genetic liability or causal effects captured by PRS and MR.

Educational attainment PRS associations was inversely associated with AD in both females and males, but the female-male difference did not survive FDR correction. This supports an association between education-related genetic liability and AD while providing limited evidence that the association differs by sex. Prior sex-stratified epidemiological studies have similarly reported mixed evidence for sex differences in the association between education and dementia ^9,38,39^. These differences across studies may reflect variation in birth cohort, social opportunity, measurement, and the meaning of educational attainment across populations. Our results therefore support education as an important AD-related pathway while providing limited evidence that its genetic association with AD differs consistently by sex.

Physical activity and social isolation showed FDR-significant female-male differences in genetic correlation without corresponding FDR-significant PRS or MR sex differences. This pattern indicates sex differences in genome-wide genetic overlap with AD but does not establish that modifying these exposures would have difference effects in females and males ^40^. These findings indicate sex differences in the shared genetic architecture of physical activity, social isolation, and AD, but do not establish sex-specific causal effects of either exposure.

Lipid and vascular traits showed a similar pattern, with sex differences in genetic correlation but limited evidence of sex heterogeneity from PRS and MR. LDL cholesterol showed an FDR-significant sex difference in genetic correlation, with positive genetic overlap with AD among males but little evidence of overlap among females. Pulse pressure similarly showed an FDR-significant female-male difference, with genetic correlations in opposite direction across sexes. Recent longitudinal vascular studies have reported sex differences in associations between blood pressure measures and cognitive decline or dementia ^41^. In our analyses, diastolic blood-pressure PRS was inversely associated with AD among males, but the female-male PRS difference was not significant. The inverse direction of some genetically informed blood-pressure associations should not be interpreted as evidence that elevated blood pressure is protective. Genetic liability reflects lifelong inherited differences and may capture biology that differs from measured mid or late-life exposures while survival treatment and selection can further complicate association with late onset disease ^4^.

Discordance across cross-trait PRS, genetic correlation and MR is informative rather than necessarily contradictory. These approaches interrogate different aspects of genetic architecture: PRS captures aggregate inherited liability at the individual level, genetic correlation quantifies genome wide sharing between traits and MR evaluates potential cause or effects under instrumental variables assumptions ^10,11^. In our study, formal sex differences concentrated in genetic correlation, whereas PRS and MR more often identified association within individual strata without demonstrating that the female and male estimates themselves differed. The distinction is important in sex-stratified research because statistical significance in one sex and not the other is not itself evidence of a sex difference ^35,41^.

The composite PRS and HABS-HD biomarker analysis provided additional context across levels of AD-related biology. The composite score was associated with AD across weighing approaches, although leave-one-PRS-out analyses showed that educational attainment contributed substantially to this signal. In HABS-HD, type 2 diabetes PRS was associated with higher Nfl, BMI PRS with higher pTau_181_ and the composite PRS with higher NfL among males after FDR correction. The limited number of biomarker associations may reflect lower power and differences between cross-sectional biomarkers and the processes represented by lifelong genetic liability.

Several limitations should guide interpretation. Sex stratification reduces effective sample size, exposure to GWAS differed in phenotype definition and statistical power, and genetic liability is not equivalent to measured exposure or intervention, and MR remains sensitive to instrument strength, heterogeneity, and horizontal pleiotropy ^11,42^. In addition, much of the available GWAS evidence remains disproportionately derived from European-ancestry populations, limiting generalizability ^14^. This is particularly important for modifiable risk factors because both genetic architecture and the social and environmental contexts in which these traits are expressed vary across population ^1,5,9^. Consequently, sex specific genetic estimate derived from one ancestry or social context should not automatically be assumed to generalize to others.

Overall, most female-male comparisons did not remain significant after FDR correction, and PRS and MR provided little evidence of sex heterogeneity. The exceptions were genetic correlations involving social isolation, physical activity, LDL cholesterol, and pulse pressure. Because these correlations describe shared genetic architecture rather than causal effects, they do not support sex-specific prevention recommendations. Replication in larger, ancestrally diverse, sex-stratified studies is needed to determine the relevance of these signals.

## Data Availability

Data for ADGC and HABS-HD are available from https://www.adgenetics.org/ and https://healthandagingbrainstudy.org/, respectively. Information about the GWAS Summary statistics used for each trait is provided in Supplementary Table S1.

https://healthandagingbrainstudy.org/

## ACKNOWLEDGEMENTS

**Alzheimer’s Disease Genetics Consortium (ADGC)**

Data for this study were prepared, archived, and distributed by the National Institute on Aging Alzheimer’s Disease Data Storage Site (NIAGADS) at the University of Pennsylvania (U24-AG041689), funded by the National Institute on Aging.

The Alzheimer’s Disease Genetics Consortium (ADGC) supported sample preparation, sequencing and data processing through NIA grant U01AG032984. Sequencing data generation and harmonization is supported by the Genome Center for Alzheimer’s Disease, U54AG052427, and data sharing is supported by NIAGADS, U24AG041689. Samples from the National Centralized Repository for Alzheimer’s Disease and Related Dementias (NCRAD), which receives government support under a cooperative agreement grant (U24 AG021886) awarded by the National Institute on Aging (NIA), were used in this study. We thank contributors who collected samples used in this study, as well as patients and their families, whose help and participation made this work possible.

Research reported in this publication was supported by the National Institute on Aging of the National Institutes of Health under Award Numbers R01AG054073 and R01AG058533, R01AG070862, P41EB015922 and U19AG078109. The content is solely the responsibility of the authors and does not necessarily represent the official views of the National Institutes of Health. We gratefully acknowledge the contributions of our study partners and their families, whose help and participation made this work possible.

**Health & Aging Brain Study—Health Disparities (HABS-HD)**

The authors would like to thank all the participants, staff, researcher teams, and partners of the Health & Aging Brain Study—Health Disparities (HABS-HD). The HABS-HD is supported by the National Institute on Aging of the National Institutes of Health under Award Numbers R01AG054073, R01AG058533, R01AG070862, P41EB015922, and U19AG078109. The content is solely the responsibility of the authors and does not necessarily represent the official views of the National Institutes of Health. We gratefully acknowledge the contributions of our study partners and their families, whose help and participation made this work possible.

^†^HABS-HD Study Team membership: MPIs-Sid E O’Bryant, Kristine Yaffe, Arthur Toga, Robert Rissman, & Leigh Johnson; and the HABS-HD Investigators: Meredith Braskie, Kevin King, James R Hall, Melissa Petersen, Raymond Palmer, Robert Barber, Yonggang Shi, Fan Zhang, Rajesh Nandy, Roderick McColl, David Mason, Bradley Christian, Nicole Phillips, Stephanie Large, Joe Lee, Badri Vardarajan, Monica Rivera Mindt, Amrita Cheema, Lisa Barnes, Mark Mapstone, Annie Cohen, Amy Kind, Ozioma Okonkwo, Raul Vintimilla, Zhengyang Zhou, Michael Donohue, Rema Raman, Matthew Borzage, Michelle Mielke, Beau Ances, Ganesh Babulal, Jorge Llibre-Guerra, Carl Hill and Rocky Vig.

## Data availability

Data for ADGC and HABS-HD is available from https://www.adgenetics.org/ and https://healthandagingbrainstudy.org/ respectively. Information about the GWAS Summary statistics used for each trait is provided in Supplementary Table S1.

## Code availability

https://github.com/AndrewsLabUCSF/sex-differences-ad-genetic-risk

