## Supplementary Methods for "Sex differences in genetic liability to modifiable Alzheimer’s disease risk factors"

### **S1. Preprocessing and Quality Control of the ADGC Genotype Dataset**

Alzheimer’s Disease Genetics Consortium (ADGC) genotype data underwent standard preprocessing and quality control at both the sample and variant levels. Pre-imputation quality control was performed using PLINK v1.9, excluding variants with high missingness, low call rates, Hardy–Weinberg equilibrium (HWE) deviation, or differential missingness between cases and controls. Samples were also excluded for low genotype call rates, sex discordance based on X-chromosome heterozygosity, or ancestry misclassification based on comparison with 1000 Genomes Phase 3 reference populations. Relatedness and population structure were assessed using approximately 20,000 linkage-disequilibrium (LD)-pruned variants, with ancestry outliers excluded based on principal component analysis (>6 standard deviations from the expected ancestry cluster). Genetic principal components were retained as covariates in downstream analyses.

ADGC genotype datasets were imputed to the Trans-Omics for Precision Medicine (TOPMed) R5 haplotype reference panel using the TOPMed Imputation Server, with EAGLE for phasing and Minimac4 for imputation. The TOPMed reference panel contains more than 300 million unique variants derived from a large and ancestrally diverse sample. Variants were mapped to the GRCh38 genome build.

Post-imputation quality control of the merged ADGC genotype dataset was performed using PLINK2 and GenoTools. The merged dataset initially contained 75,886 individuals and 5,300,619 variants. Samples with >5% missing genotype calls were excluded (--mind 0.05; n = 4,801), leaving 71,085 individuals, including 42,305 females, 28,761 males, and 19 individuals with ambiguous sex.

For relatedness estimation, variants showing extreme deviation from HWE (P < 1 × 10^-30) or minor allele count <2 were excluded. The HWE filter removed 1,102,155 variants, whereas no additional variants were removed by the minor allele count threshold, leaving 4,198,464 variants for kinship estimation. Pairwise kinship coefficients were estimated using KING. A kinship coefficient ≥0.0884, corresponding approximately to second-degree relatives or closer, identified 17,585 related pairs. Relatedness pruning using this threshold reduced the dataset from 71,085 to 61,228 individuals. A higher kinship threshold of ≥0.354 was additionally evaluated to identify duplicate or monozygotic-twin relationships.

Heterozygosity was assessed using a subset of common, high-quality, approximately LD-independent variants. Variants with genotype missingness >1% or minor allele frequency <5% were excluded, followed by LD pruning using a 50-variant window, a five-variant step size, and an r² threshold of 0.5. This yielded 469,587 variants for heterozygosity estimation. Nine heterozygosity outliers were excluded, leaving 61,219 individuals.

A final HWE filter at P < 1 × 10^-30 removed an additional 1,999 variants. Following post-imputation quality control, the final dataset contained 61,219 individuals, including 36,287 females, 24,915 males, and 17 individuals with ambiguous sex, and 4,196,465 variants. Individuals with ambiguous sex were excluded from sex-stratified downstream analyses.

### **S2. ADGC Phenotype Definition and Analytic Sample**

Following genotype quality control, participants were linked to ADGC phenotype and covariate data and to ancestry-standardized polygenic risk scores (PRSs). Alzheimer’s disease (AD) status was defined using the harmonized ADGC clinical phenotype. Participants classified as affected with AD were designated as cases, and cognitively unimpaired participants were designated as controls. Participants with mild cognitive impairment, missing diagnosis, or other diagnostic categories were excluded from the primary case-control analysis.

The PRS analytic sample was restricted to participants with non-missing AD status; age at onset for AD cases or age at examination for controls; *APOE e4* carrier status; sex; and the first four genetic principal components. Participants were additionally required to have ancestry-standardized scores available for all 17 modifiable-risk-factor PRSs included in the analysis. Following application of these phenotype, covariate, and PRS availability criteria, 27,751 participants were included in the primary ADGC PRS analysis.

Sex-stratified analyses were conducted separately among participants classified as female or male. Individuals with ambiguous or missing sex were excluded from these analyses.

### **S3. HABS-HD Phenotype Definitions and Analytic Samples**

The Health and Aging Brain Study–Health Disparities (HABS-HD) is an ongoing community-based longitudinal study designed to investigate biological, medical, behavioral, and social determinants of brain aging and Alzheimer’s disease and related dementias in a racially and ethnically diverse population. HABS-HD includes non-Hispanic Black, Hispanic, and non-Hispanic White adults and incorporates detailed clinical, cognitive, demographic, medical, social, genetic, and biomarker characterization.

Genotype data from HABS-HD releases were merged using PLINK. The merged genotype dataset contained 3,314 participants and 650,181 variants. Participants were subsequently linked to phenotype, covariate, and polygenic risk score data for the present analyses.

Demographic characteristics, including age, sex, self-reported race and ethnicity, education, and language, as well as medical history and social determinants of health, were collected using standardized clinical interviews and electronic data-capture procedures. For the present study, HABS-HD variables were selected to correspond as closely as possible to the phenotypes represented by the GWAS used to construct each risk-factor PRS.

Matched phenotypes included continuous or binary measures corresponding to educational attainment, alcohol consumption, smoking, body mass index, type 2 diabetes, systolic blood pressure, diastolic blood pressure, pulse pressure, HDL cholesterol, LDL cholesterol, total cholesterol, triglycerides, depression, social isolation or social support, physical activity, hearing loss, and vision loss, according to available HABS-HD measures.

The primary HABS-HD analytic sample included 3,207 participants after linkage of genetic, phenotype, and covariate data. Analytic sample sizes varied across individual phenotype-validation and biomarker analyses according to availability of the corresponding phenotype or biomarker and required covariates.

### **S4. GWAS Summary Statistics and Polygenic Risk Score Construction**

Published genome-wide association study (GWAS) summary statistics were obtained for 17 modifiable dementia risk factors: educational attainment, alcohol consumption, smoking initiation, BMI, type 2 diabetes, systolic blood pressure, diastolic blood pressure, pulse pressure, HDL cholesterol, LDL cholesterol, total cholesterol, triglycerides, depression, social isolation, physical activity, hearing loss, and vision loss. GWAS source, phenotype definition, ancestry, sample size, genome build, and use in each downstream analysis are provided in Supplementary Table S1.

Summary statistics were standardized and harmonized before PRS construction. Polygenic scores were generated using PRS-CS Auto, which uses a Bayesian continuous-shrinkage prior to estimate posterior SNP effect sizes while accounting for LD. Analyses were restricted to HapMap3 variants available in the relevant LD reference panel and target genotype dataset.

Posterior SNP weights estimated by PRS-CS Auto were applied to individual-level genotype data to generate trait-specific PRSs. Scores were subsequently processed using pgsc_calc to obtain ancestry-standardized PRSs for downstream analyses. Each ancestry-standardized PRS was standardized to mean 0 and standard deviation 1 before association testing, so that reported effect estimates corresponded to a one-standard-deviation increase in genetic liability to the corresponding trait.

The original GWAS effect direction was retained for analyses of individual PRSs so that higher PRS values represented greater genetically predicted levels or liability according to the phenotype definition of the source GWAS.

### **S5. Composite Polygenic Risk Score**

We constructed a multi-trait composite PRS to evaluate the cumulative contribution of genetic liability across modifiable dementia risk-factor domains to AD case–control status. For composite-score construction, component PRSs were directionally aligned so that higher values consistently represented the prespecified direction of greater dementia-risk liability. PRSs for traits whose original GWAS direction represented a putatively protective phenotype were sign-reversed for composite-score construction only. Risk-factor orientation used in reporting is summarized in Supplementary Table S3.

The ADGC analytic sample was divided into a 70% training set and a 30% held-out test set using stratified random sampling based on AD case–control status. Female- and male-specific training and test datasets were generated from the corresponding partitions for sex-stratified analyses.

Within each training set, component PRSs were standardized using the training-set mean and standard deviation. The resulting scaling parameters were then applied unchanged to the corresponding held-out test set to avoid information leakage.

For the primary unpenalized composite model, all standardized component PRSs were entered simultaneously into a multivariable logistic regression model fitted in the training set. Models were adjusted for age, *APOE e4* carrier status, the first four genetic principal components, and sex in the combined-sex analysis. Sex was omitted from sex-stratified models. Regression coefficients corresponding to the component PRSs were then used as weights to construct an individual-level composite PRS.

The resulting composite score was standardized using its standard deviation in the training set, and the same scaling factor was applied to the held-out test set. Associations between the composite PRS and AD case–control status were evaluated exclusively in the held-out test set using logistic regression with the same covariate structure as the corresponding training model.

Because several component PRSs were correlated, ridge and elastic-net logistic regression models were evaluated as sensitivity analyses. Penalized models were trained using 10-fold cross-validation within the training dataset. The penalty parameter λ was selected using the value minimizing cross-validated prediction error. Ridge regression used α = 0, and elastic-net regression used α = 0.5. Standardized component PRSs and model covariates were included in the penalized models, and coefficients corresponding to the PRS terms were used as weights to construct penalized composite scores. These scores were standardized using parameters derived from the training dataset and evaluated in the held-out test set using the same logistic regression framework as the primary composite score.

To assess whether the composite association was disproportionately influenced by a single component PRS, we performed leave-one-PRS-out analyses. Each component PRS was sequentially removed, the unpenalized composite model was retrained using the remaining scores, and the resulting composite PRS was evaluated in the corresponding held-out test set. Effect estimates and test-set discrimination, quantified using the area under the receiver operating characteristic curve (AUC), were compared with those from the full composite model.

### **S6. PRS–Phenotype Validation in HABS-HD**

We assessed whether each trait-specific PRS captured genetic liability to its corresponding measured phenotype in HABS-HD. Each PRS was paired with the available HABS-HD phenotype most closely corresponding to the phenotype definition used in the source GWAS.

Associations were evaluated using linear regression for continuous outcomes and logistic regression for binary outcomes. Continuous outcomes were standardized before model fitting so that regression coefficients represented the change in outcome standard deviations associated with a one-standard-deviation increase in the corresponding PRS. Logistic regression coefficients represented the change in log odds of the corresponding binary phenotype per one-standard-deviation increase in PRS.

Models were adjusted for age, sex, *APOE e4* carrier status, self-reported race/ethnicity, and the first four genetic principal components. Analyses were conducted using participants with complete data for the PRS, corresponding phenotype, and required covariates. Phenotypes with very low case counts were interpreted cautiously because of limited precision.

### **S7. Plasma Biomarker Analyses in HABS-HD**

Associations between trait-specific PRSs and plasma biomarkers related to AD pathology and neurodegeneration were evaluated in HABS-HD. Biomarker outcomes included plasma phosphorylated tau_181_ (*pTau_181_*), phosphorylated tau 217 (*pTau_217_*), amyloid-β_42_/amyloid-β_40_ ratio (*Aβ_42_/Aβ_40_*), neurofilament light chain (NfL), and total tau.

Biomarker outcomes were standardized before model fitting. Associations with each standardized trait-specific PRS were evaluated using linear regression adjusted for age, sex, *APOE e4* carrier status, self-reported race/ethnicity, and the first four genetic principal components.

The composite PRS was evaluated using the same analytical framework. Sex-stratified biomarker analyses were performed separately in females and males using the corresponding covariate structure with sex omitted.

Effect estimates therefore represent the standard-deviation difference in the biomarker associated with a one-standard-deviation increase in the PRS. Analyses used complete observations for the PRS, biomarker, and model covariates.

### **S8. Individual Polygenic Risk Score Associations with Alzheimer’s Disease**

Associations between each standardized trait-specific PRS and AD case–control status were evaluated in ADGC using logistic regression. In the combined-sex analysis, models were adjusted for age, *APOE e4* carrier status, sex, and the first four genetic principal components. Analyses were then repeated separately among female and male participants, with sex omitted as a covariate.

Effect estimates were reported as odds ratios (ORs) and 95% confidence intervals corresponding to a one-standard-deviation increase in the PRS.

Differences between female- and male-specific PRS associations were evaluated using a Z-test:

Z = (β_F − β_M) / √(SE_F² + SE_M²),

where β_F and β_M represent the female- and male-specific logistic regression coefficients and SE_F and SE_M their corresponding standard errors. Two-sided P values were calculated from the resulting Z statistics.

For trait-specific PRS analyses, P values were adjusted for multiple testing using the Benjamini–Hochberg false discovery rate (FDR) procedure separately for the combined-sex, female-specific, male-specific, and female-versus-male comparison analyses. Associations with unadjusted P < 0.05 but FDR-adjusted P ≥ 0.05 were considered nominal, whereas associations with FDR-adjusted P < 0.05 were considered significant after correction for multiple testing.

### **S9. Genome-Wide Genetic Correlation**

Genome-wide genetic correlations between each modifiable dementia risk factor and AD were estimated using GNOVA. Analyses were performed separately using combined-sex, female-specific, and male-specific GWAS summary statistics. Female-specific risk-factor GWAS were paired with female-specific AD GWAS, male-specific risk-factor GWAS with male-specific AD GWAS, and combined-sex risk-factor GWAS with combined-sex AD GWAS.

Before analysis, GWAS summary statistics were standardized and harmonized, and variants were restricted to HapMap3 SNPs. The major histocompatibility complex region on chromosome 6 (26–34 Mb) was excluded because of its extended and complex LD structure. Variants with missing required information following preprocessing were removed.

GNOVA analyses were performed using European-ancestry LD reference genotype data restricted to common variants. Genetic correlation estimates (r_g_), standard errors, and P values corrected by GNOVA for potential sample overlap were retained. Exposure–outcome pairs for which genetic correlation could not be estimated because of non-positive SNP heritability were excluded. Estimates with |r_g_| >1 were also excluded from inferential analyses.

The primary analysis used AD GWAS summary statistics from a meta-analysis combining ADGC, ADSP, UK Biobank, and FinnGen. Sensitivity analyses used alternative AD GWAS meta-analyses excluding either FinnGen or UK Biobank to evaluate whether results were disproportionately influenced by either contributing cohort.

Differences between female- and male-specific genetic correlation estimates were evaluated using:

Z = (r_g_ ,F − r_g_ ,M) / √(SE_F² + SE_M²),

where r_g_,F and r_g_,M represent the female- and male-specific genetic correlation estimates and SE_F and SE_M their corresponding standard errors. Two-sided P values were calculated from the resulting Z statistics.

P values were corrected for multiple testing using the Benjamini–Hochberg FDR procedure separately within each AD GWAS source and sex stratum. P values from direct female-versus-male comparisons were FDR-adjusted separately within each AD GWAS source. Unadjusted P < 0.05 with FDR-adjusted P ≥ 0.05 was considered nominal evidence, whereas FDR-adjusted P < 0.05 was considered significant after multiple-testing correction.

### **S10. Mendelian Randomization**

Two-sample Mendelian randomization (MR) was performed to evaluate whether genetically predicted modifiable dementia risk factors were associated with AD risk. Genetic instruments for each exposure were defined as genome-wide significant variants (P < 5 × 10^-8). Independent instruments were selected using LD clumping at r² < 0.001 within a 10-Mb window.

Exposure and AD outcome summary statistics were harmonized using the TwoSampleMR package. Effect alleles were aligned between exposure and outcome datasets before analysis. Variants associated with the AD outcome at genome-wide significance (P ≤ 5 × 10^-8) were excluded to reduce the potential influence of variants with strong direct associations with the outcome. Variants within the APOE region (GRCh37 chromosome 19:44,909,053–45,912,650) were additionally excluded. When an exposure instrument was unavailable in the outcome GWAS, proxy variants in LD (r² ≥ 0.8 within 500 kb) were used when available.

MR analyses were restricted to exposure–outcome pairs with at least three retained genetic instruments. The primary causal estimate was obtained using fixed-effect inverse-variance weighted (IVW) MR. Weighted-median, weighted-mode, and MR-Egger regression were performed as complementary sensitivity analyses.

Between-instrument heterogeneity was assessed using Cochran’s Q statistic. Directional horizontal pleiotropy was evaluated using the MR-Egger intercept. I²_GX was calculated to assess potential regression dilution and instrument-strength heterogeneity in MR-Egger analyses. MR-PRESSO was additionally used to identify potential outlier instruments. When outliers were identified, the relevant MR analyses were repeated following removal of the detected outlier variants.

Analyses were conducted separately using combined-sex, female-specific, and male-specific exposure and AD GWAS summary statistics, with sex-stratified exposure GWAS paired with the corresponding sex-stratified AD GWAS.

Differences between female- and male-specific IVW estimates were evaluated using:

Z = (β_F − β_M) / √(SE_F² + SE_M²),

where β_F and β_M represent the female- and male-specific IVW estimates and SE_F and SE_M their corresponding standard errors. This test treats the female- and male-specific effect estimates as independent. Two-sided P values were calculated from the resulting Z statistics.

For the primary IVW analyses, P values were corrected for multiple testing using the Benjamini–Hochberg FDR procedure separately within each AD outcome and sex stratum. P values from the female-versus-male comparison were FDR-corrected separately within each AD outcome. An unadjusted P < 0.05 with FDR-adjusted P ≥ 0.05 was considered nominal evidence, whereas FDR-adjusted P < 0.05 was considered significant after multiple-testing correction.

The primary AD outcome was the GWAS meta-analysis combining ADGC, ADSP, UK Biobank, and FinnGen. Sensitivity analyses were conducted using alternative meta-analyses comprising ADGC, ADSP, and UK Biobank, thereby excluding FinnGen, and ADGC, ADSP, and FinnGen, thereby excluding UK Biobank. Primary IVW findings were interpreted together with weighted-median, weighted-mode, MR-Egger, heterogeneity, pleiotropy, and MR-PRESSO results rather than on the basis of the IVW estimate alone.

**SUPPLEMENTARY FIGURES**
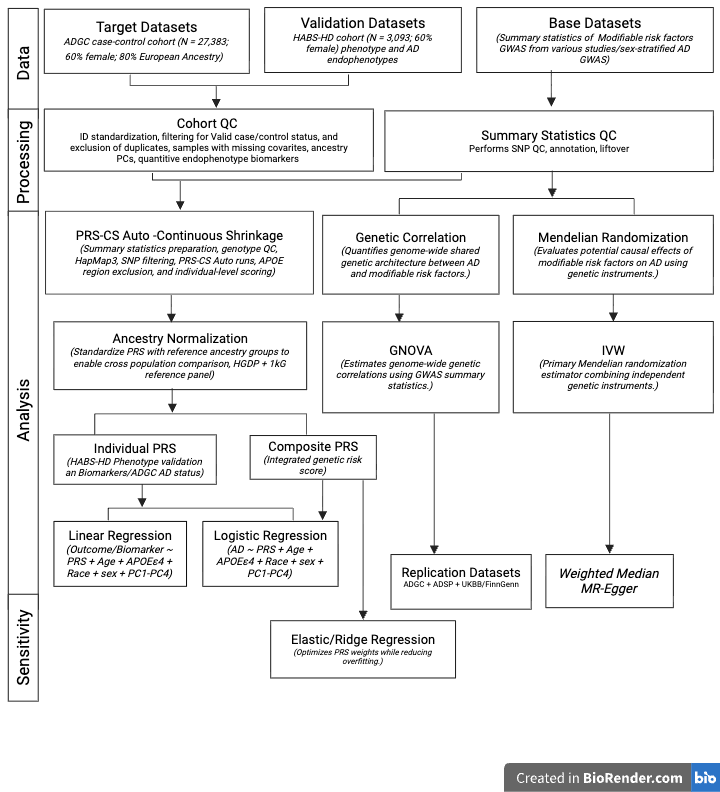
 **Supplementary Figure S1. Overview of the study design and analytic workflow.**

The study integrated polygenic risk score (PRS), genome-wide genetic correlation, and two-sample Mendelian randomization (MR) analyses to investigate the relationship between genetic liability to modifiable dementia risk factors and Alzheimer’s disease (AD). Individual and composite PRSs were constructed from published genome-wide association study (GWAS) summary statistics following quality control and ancestry normalization. PRSs were evaluated in the Alzheimer’s Disease Genetics Consortium (ADGC) for associations with AD case–control status and in the Health and Aging Brain Study–Health Disparities (HABS-HD) for phenotype validation and associations with AD-related plasma biomarkers. Genome-wide shared genetic architecture between AD and modifiable risk factors was assessed using GNOVA to estimate genetic correlation (*r*g). Potential causal relationships were evaluated using inverse-variance weighted (IVW) Mendelian randomization, with weighted median and MR-Egger methods performed as complementary sensitivity analyses. Composite PRSs were constructed using penalized regression. Unless otherwise specified, regression models were adjusted for age, *APOE e4* status, genetic principal components, and sex (overall analyses), with additional adjustment for self-reported race/ethnicity in HABS-HD.

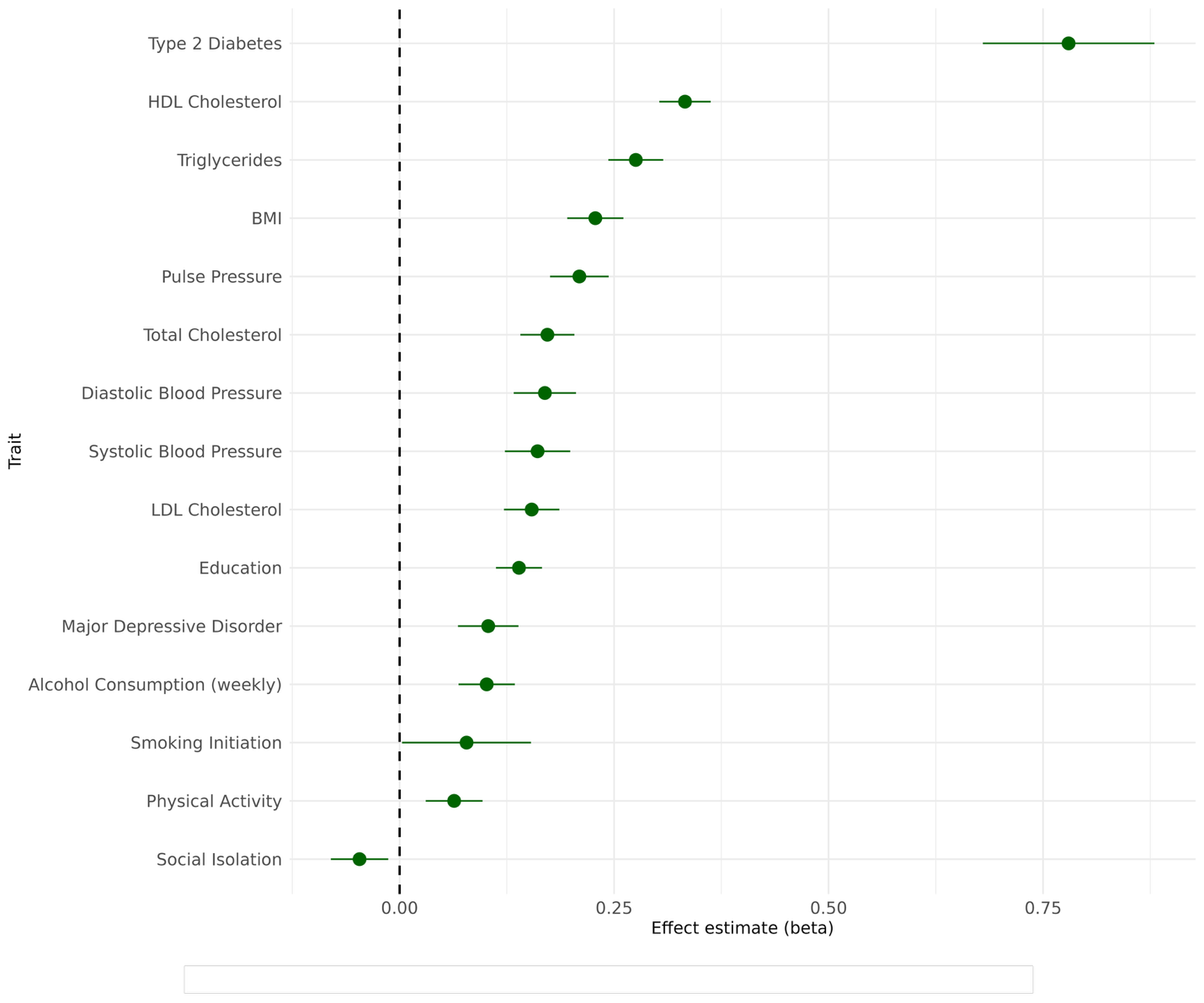

**Supplementary Figure S2**. **Phenotype validation of individual polygenic risk scores (PRSs) in HABS-HD.** Points show effect estimates for the association between each standardized PRS and its matched phenotype in the HABS-HD analytic sample, adjusted for age, *APOE e4* status, self-reported race/ethnicity, and genetic principal components (PC1–PC4). Error bars indicate 95% confidence intervals. Continuous phenotypes are presented as standardized beta coefficients, and binary phenotypes are presented as log-odds beta coefficients from logistic regression. The dashed vertical line marks the null value of 0. Traits are ordered by effect estimate. Positive effect estimates indicate that higher PRS is associated with higher values or greater odds of the corresponding phenotype. Vision loss and hearing loss were excluded from the phenotype-validation plot because the observed case counts in HABS-HD were very low, making those matched-phenotype estimates unstable.

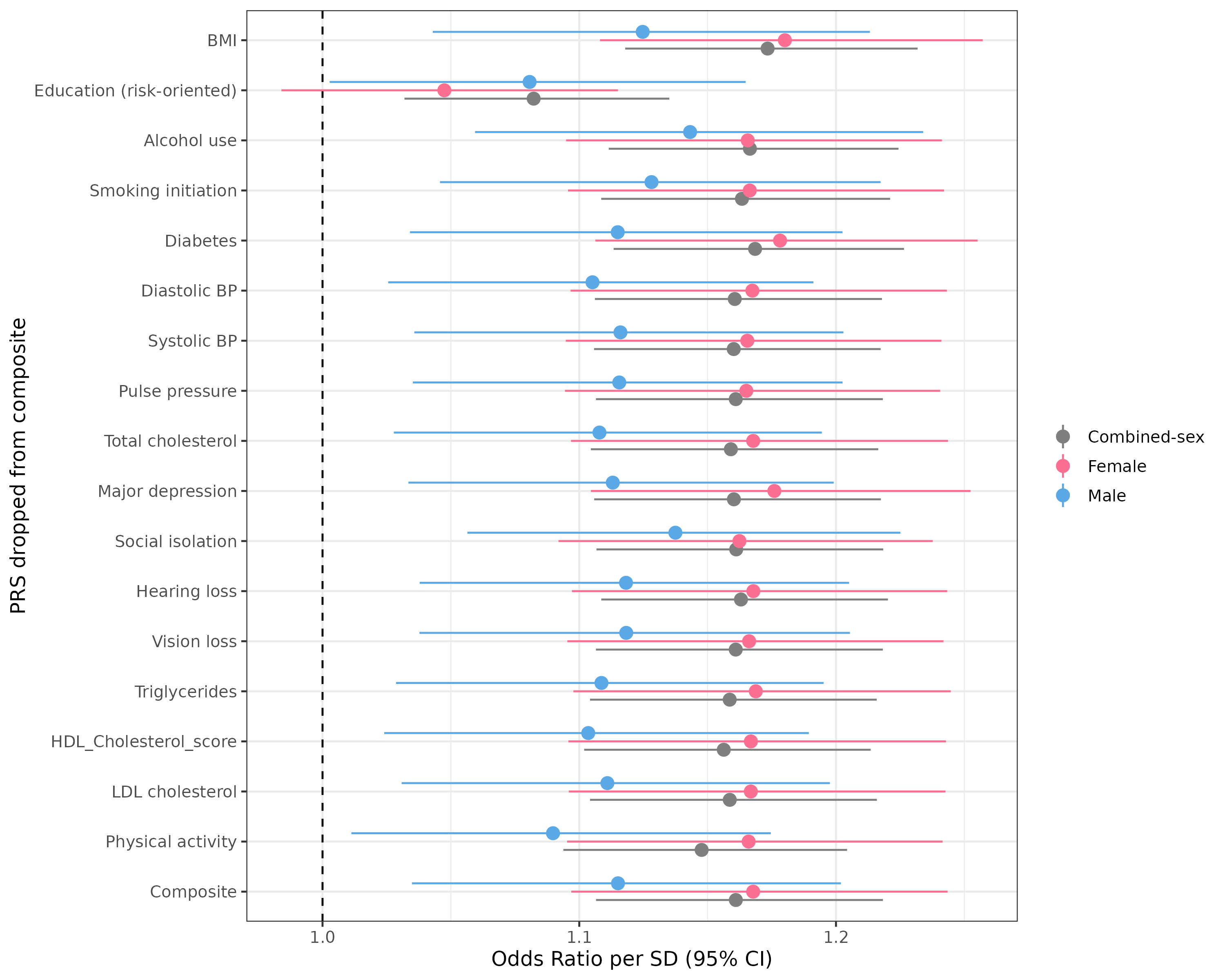

**Supplementary Figure S3. Leave-one-PRS-out composite PRS analysis in ADGC**. Odds ratios (ORs) and 95% confidence intervals are shown for associations between each leave-one-PRS-out composite score and Alzheimer’s disease case–control status in the combined-sex (gray), female-specific (pink), and male-specific (blue) held-out test sets. Each row identifies the component PRS omitted when the composite score was retrained; the full composite score is shown for comparison. Composite scores were constructed separately in the corresponding training samples and evaluated in held-out test samples. Models were adjusted for age, *APOE e4* carrier status, and the first four genetic principal components; sex was additionally included in the combined-sex model. The dashed vertical line denotes the null value of OR = 1.

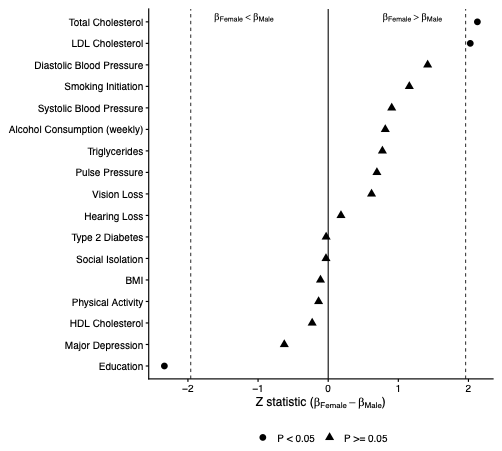

**Supplementary Figure S4: Female-male differences in associations between individual polygenic risk squares and Alzheimer's disease in ADGC.** The Z statistic was calculated as *Z = (β_F_ - β_M_)* / *√((SE_F_)^2^* *+ (SE_M_)^2^),* Using logs odds coefficient from sex stratified logistic regression models. Positive Z values indicate *β_F_*  > *β_M_,* whereas negative values indicate *β_F_* < *β_M_.* The solid vertical line denotes no female male difference (Z = 0), and the dashed vertical lines to note Z = ± 1.96, Corresponding to a nominal two-sided P < 0.05. Circles indicate nominally significant female-male differences (P < 0.05), and triangles indicate P ≥ 0.05). Models were adjusted for age, *APOE ε4* status and the first four genetic ancestry principal components.

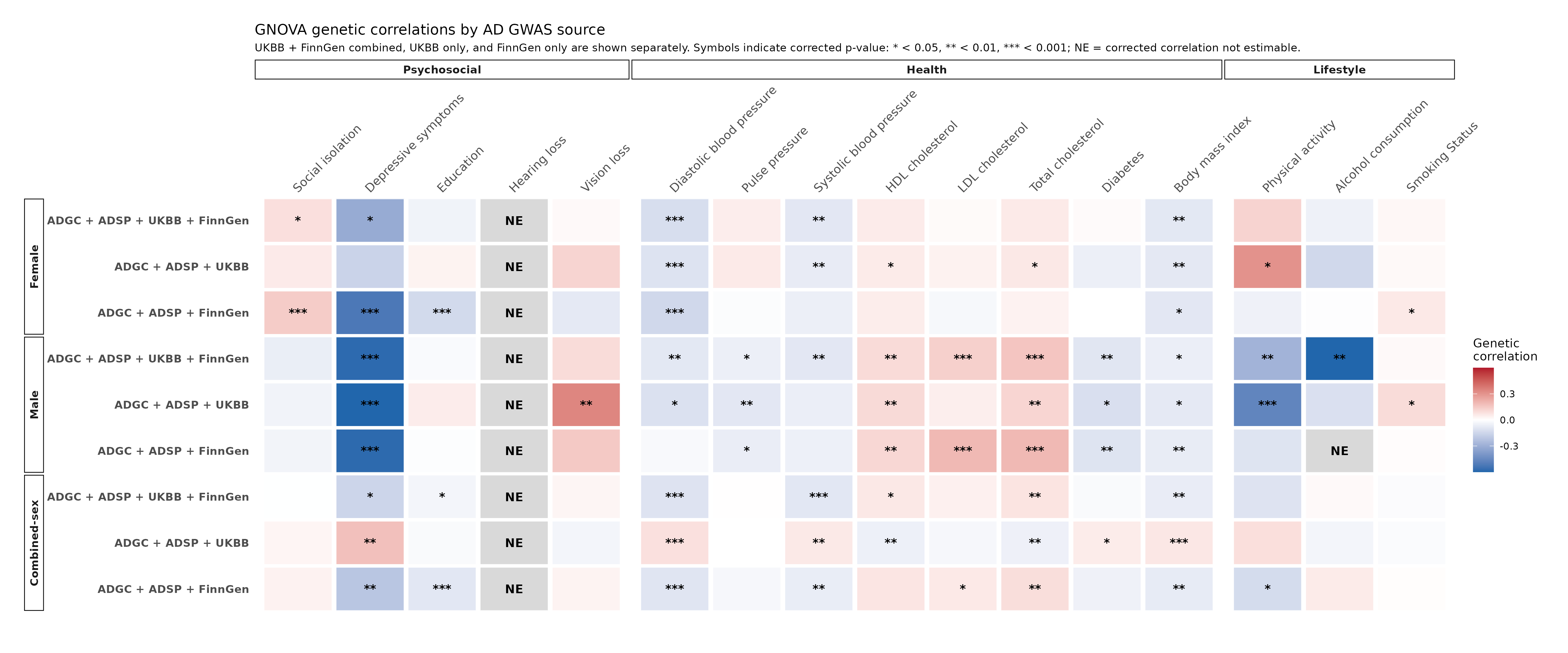

**Supplementary Figure S5. Genome-wide genetic correlations between modifiable dementia risk factors and Alzheimer’s diseases across AD GWAS sources.** Heatmap showing GNOVA genetic-correlation estimates from female-specific, male-specific, and combined-sex analyses. Within Each six stratum, row represents the primary AD GWAS meta-analysis combining ADGC, ADSP, UK Biobank (UKBB), and FinnGen; a sensitivity analysis excluding FinnGen (ADGC, ADSP, UKBB); and a sensitivity analysis excluding UKBB (ADGC, ADSP, FinnGen). Columns represent modifiable dementia risk factors grouped in psychosocial, health, and lifestyle domains. Effect directions were harmonized to represent greater dementia risk liability; educational attainment, HDL cholesterol, and physical activity were therefore oriented as lower education attainment, lower HDL cholesterol, and lower physical activity respectively. Red indicates a positive genetic-correlation, and blue indicates an inverse genetic-correlation. Asterisks indicate association surviving FDR correction with each analysis stratum (*q < 0.05, q < 0.01, q < 0.001).* Asterix refer to the significance of the genetic correlation estimate and not the female-male difference test. NE indicates that the genetic correlation could not be estimated. Complete estimates, standard errors, nominal P values, and FTR adjusted P values are provided in Supplementary Table S7.

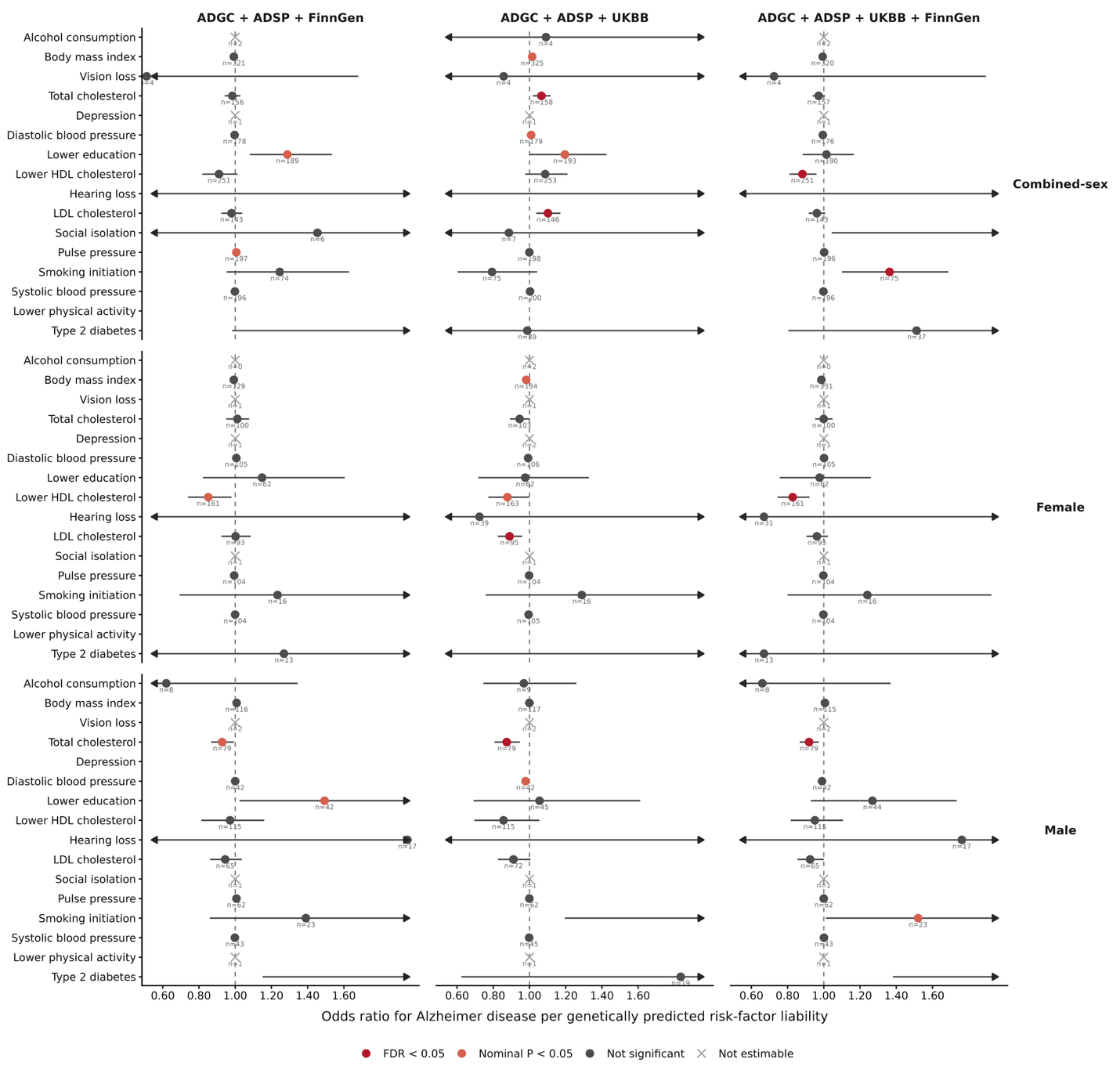

**Supplementary Figure S6. Mendelian Randomization estimates for modifiable dementia risk factors across AD GWAS outcomes.** Fixed effect-inverse-variance-weighted estimates are shown for combined-sex, female-specific and male-specific analysis using three AD outcome GWAS definitions: ADGC + ADSP + FinnGen, ADGC + ADSP + UKBB, and ADGC + ADSP + FinnGen + UKBB. Points denote odd odds ratio for AD per genetically predicted risk factor liability, and horizontal line denotes 95% confidence intervas;l arrows indicate confidence intervals extending beyond the plotting range. Effects were oriented so that an odds ratio greater than 1 indicate that greater genetic genetically predicted risk-factor liability is associated with greater AD risk. Red points indicate FDR-adjusted P < 0.05, orange points indicate nominal P < 0.05 but FDR adjusted P ≥ 0.05, gray points indicate non-significant estimates, and crosses indicates estimates that were not estimable. Numbers adjacent to the estimates indicate the numbers of restrained genetic instruments.
